# Longitudinal trajectories of global and domain-specific cognition after stroke using the Oxford Cognitive Screen

**DOI:** 10.1101/2025.11.14.25340284

**Authors:** Elise Milosevich, Andrea Kusec, Sarah T. Pendlebury, Nele Demeyere

## Abstract

**Background:** Cognitive impairment is common after stroke and linked to poor outcomes, yet long-term recovery or decline, particularly across specific cognitive domains, remains unclear. Most studies use brief global screeners with short follow-up, limiting insight into recovery patterns. This study aimed to characterize domain-specific cognitive trajectories over ≥2 years post-stroke and identify predictors of persistent impairment.

**Methods:** Participants were assessed acutely, at 6 months, and ≥2 years post-stroke (Median = 4.1 years [IQR = 3.3]). The Oxford Cognitive Screen (OCS) was administered at all timepoints. Global impairment severity was quantified by the proportion of OCS subtasks impaired. Logistic mixed-effects models examined longitudinal change and predictors of domain-specific impairments (language, memory, attention, executive function, number processing). Latent class growth analysis (LCGA) identified distinct cognitive trajectories.

**Results:** Of 105 patients assessed, 98 had complete OCS data. Cognitive impairment severity improved substantially by 6 months (β = −0.11) and further long term (>2 years; β = −0.15). Acute impairment severity strongly predicted long-term outcomes (β = 0.50; p < 0.001), while demographic and vascular factors explained minimal variance. LCGA identified four overall trajectories: no or mild acute impairment with stability (47.6%), moderate-improving (32.3%), large improvement (15.2%), and decline (4.8%). Domain-specific improvements were greatest in memory (OR = 16.40) and language (OR = 8.17), more limited in attention (OR = 5.41) and executive function (OR = 4.14). Domain models revealed additional classes of persistent or delayed recovery, particularly in executive function and attention.

**Conclusions:** Cognitive recovery is most pronounced within 6 months and continues across domains, though executive dysfunction often persists. Acute impairment severity best predicted long-term outcomes, while vascular and demographic factors were less informative. Distinct trajectory classes highlight the need for individualized, long-term cognitive monitoring to guide rehabilitation and prognostication. These findings underscore the importance of long-term cognitive follow-up in stroke care and provide empirical benchmarks for recovery across domains.

## INTRODUCTION

Cognitive impairment is common following stroke (1, 2)^i^ and frequently contributes to poor functional outcomes (3, 4), increased dependency (5), and reduced quality of life (4). While many individuals experience some degree of cognitive recovery, others may show persistent impairment or delayed decline (3, 6). However, the long-term trajectories of change across different cognitive domains remain poorly understood (7). This is partly due to cognitive deficits often being overlooked beyond the immediate post-acute period unless dementia develops, which is not an inevitable outcome (6, 8, 9).

Most longitudinal studies of post-stroke cognition have assessed performance at only one or two timepoints within the first year (10) often using brief global screening tools or undifferentiated assessments (6). While these tools offer a general overview, they lack the sensitivity to detect domain-specific patterns of impairment and recovery that are common following stroke (11). Furthermore, cognition is frequently conceptualised dichotomously, either impaired or unimpaired, masking within-person variability and limiting insight into the heterogeneity of outcomes. Even large-scale efforts to map long-term trajectories, such as the recent pooled analysis of over 20,000 individuals across 14 cohorts (12), found consistent evidence of acute and accelerated post-stroke cognitive decline, but were constrained by variability in cognitive measures and limited resolution of domain-specific change. Consequently, it remains unclear whether change in specific cognitive domains may occur in later years, as observed in scarce longer-term (10-year) follow-up studies (8, 13).

Determining which cognitive domains are most susceptible to long-term decline, and which early impairments persist, resolve, or fluctuate, is essential for improving prognostic accuracy and informing long-term care planning (14), and extending the care pathway to cognitive monitoring (15). While several risk factors for post-stroke cognitive impairment (PSCI) have been identified, such as age, stroke severity, education level, and vascular comorbidities (9, 16), a recent systematic review highlighted the most predictive factor to be baseline cognition (17). However, few studies have investigated how this relates to domain-specific cognitive trajectories over extended periods.

To address the critical knowledge gap around longer-term domain specific cognitive trajectories after stroke, this study aimed to delineate long-term trajectories of global and domain-specific cognitive function following stroke, and to examine how early cognitive impairments, clinical variables, and demographic factors relate to different patterns of cognitive recovery or decline.

## METHODS

### Participants

Participants were initially recruited through the Oxford Cognitive Screening Programme based within the regional acute stroke unit at the John Radcliffe Hospital, UK, from 2012 to 2019 (National Research Ethics Committee (UK) approval references 14/LO/0648 and 18/SC/0550). Participants were included if they had a confirmed stroke diagnosis, aged ≥18 years and had sufficient English language comprehension to understand assessment instructions. Patients were excluded if they were unable to provide written or witnessed informed consent, or unable to concentrate for 20 minutes as judged by the multidisciplinary team. A total of 866 stroke patients were recruited and received cognitive screening acutely (≤2 weeks post-admission; Timepoint 1 [T1]), 430 (49.7%) of whom completed 6-month follow-up (T2). Following this, 208 individuals consented to being re-contacted for potential inclusion in further research, and were invited to participate in the OX-CHRONIC long-term follow-up study (UK REC Ref: 19/SC/0520).

A total of 105 participants were enrolled in OX-CHRONIC, 98 of whom completed cognitive assessments ≥2 years post-stroke (T3), which formed the present analysis sample. Detailed recruitment and attrition across all timepoints are summarized in Figure 1. Patient demographics, comorbidities, vascular risk factors, stroke characteristics, neuroimaging and laboratory data were collected on admission or retrospectively through electronic medical records. Further information outlining OX-CHRONIC study design was published previously (18, 19) together with detailed data on attrition at 6-months (2) and beyond (OX-CHRONIC)(19).

**Figure 1.**
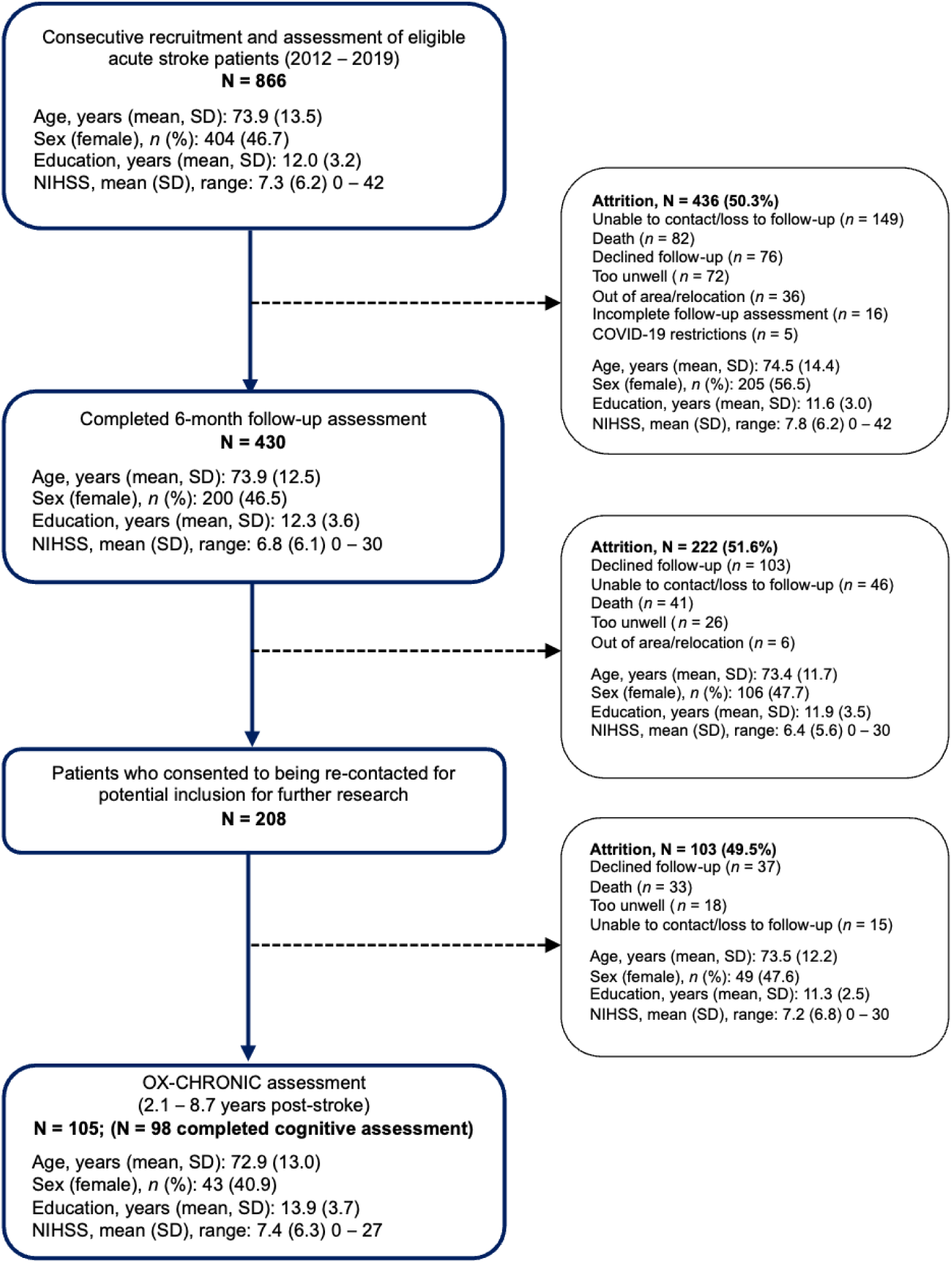
Flow-chart of participant recruitment and attrition across all assessment timepoints at acute, 6 months, ≥2 years post-stroke (chronic).

### Cognitive assessment

At all three timepoints, each participant received domain-specific cognitive screening with the Oxford Cognitive Screen (OCS)(20). The 12 OCS subtest scores were categorized into five cognitive domains: language (picture naming, semantic understanding, sentence reading), attention (egocentric and sustained spatial attention, allocentric attention), executive function (trail-making), memory (orientation, verbal recall and recognition, episodic recognition), and number processing (calculations and number writing). Subtests were binarized into impaired or unimpaired based on published normative scores for each subtest (18, 20). A domain impairment was characterized as at least one impaired subtest in that domain, as number of subtests range from 1-3 across domains. Due to COVID-19 disruption, the third timepoint (OX-CHRONIC) assessment was performed remotely through videoconferencing or telephone calls. A testing pack was mailed to the participant and the assessor provided instructions throughout the call, asking for verbal (or non-verbal, e.g., tapping phone, completing paper and pencil tasks in workbook) responses to tasks. This remote testing methodology using the OCS has since been validated (21) and remote neuropsychological assessment more generally has been found to be reliable (22). Due to the remote nature of assessment, the OCS praxis domain (gesture imitation) could not be tested, and therefore only five cognitive domains were assessed within those assessed at ≥2 years post-stroke: language, attention, executive function, memory, and number processing. The Montreal Cognitive Assessment (MoCA) (23) was used to assess general cognitive status at long-term follow-up (T3) to increase comparability of cognitive function with other studies.

### Statistical analysis

All analyses were conducted using R version 4.4.0. Descriptive statistics were used to summarize sample demographics and clinical characteristics, and to report prevalence rates of global and domain-specific impairments at each timepoint (acute, 6 months, and chronic follow-up).

To examine longitudinal trajectories of cognitive function over time, a three-stage analytic approach was applied across global and domain-specific outcomes of the 105 participants completing OX-CHRONIC assessments.

First, global cognitive function was analyzed using linear mixed-effects models, with the proportion of OCS subtasks impaired (severity of impairment) as the outcome. Fixed effects included timepoint (acute, 6 months, chronic), and random intercepts were included to account for within-subject clustering. Model 1 included time only; Model 2 added the proportion of acute OCS impairments; and Model 3 further adjusted for age, sex, education, acute NIHSS score, atrial fibrillation, hypertension, diabetes, smoking status, stroke recurrence, and days post-stroke. Models were fit using maximum likelihood estimation, with cases containing missing data excluded. Model performance was evaluated using marginal and conditional R², intraclass correlation coefficients (ICC), and fit information criteria (AIC, BIC). Pairwise comparisons between timepoints were assessed using Tukey-adjusted post-hoc tests.

Second, domain-specific impairment was modelled using mixed-effects logistic regression. For each domain (language, memory, attention, executive function, and number processing), impairment was defined as at least one subtask impaired (0 = at least one subtask impaired, 1 = unimpaired). Models included fixed effects for time and baseline impairment status (0=impaired, 1 = unimpaired), with random intercepts per participant. Model performance was reported using marginal and conditional R² and ICC. Odds ratios (ORs) and 95% confidence intervals (CIs) were calculated for all fixed effects, and Wald z-tests were used to evaluate statistical significance, with ORs signifying likelihood of cognitive recovery.

Third, to identify distinct subgroups of cognitive trajectories over time, Latent Class Growth Analysis (LCGA) was conducted using the *lcmm* package (24). Separate models were fitted for global cognition and each cognitive domain. For global cognition and the domains of language, memory, attention, and number processing, the outcome was the proportion of OCS subtasks impaired at each timepoint to allow for greater sensitivity in detecting classes. For executive function, raw mixed trail-making task scores were used. Models specifying 1 to 5 latent classes were fitted, with time as a fixed effect. Model fit was evaluated using standard information criteria, including the Akaike Information Criterion (AIC; a measure of model fit that penalizes for greater number of parameters), Bayesian Information Criterion (BIC; fit measure which penalizes number of parameters more heavily), sample-adjusted BIC (SABIC), and log-likelihood. Final model selection was guided by a combination of statistical fit, parsimony, and clinical interpretability of the identified trajectories, with entropy (H; measure of class separation) values ≥0.80 considered indicative of acceptable classification certainty. For each selected model, trajectory class membership was compared across demographic (age, sex, education), clinical (NIHSS, stroke type, recurrence, lesion hemisphere), and neuroimaging variables (lesion volume, white matter hyperintensities, cortical atrophy), as well as acute and chronic cognitive functioning (number of acute domains impaired, MoCA score at follow-up). We additionally explored between-group statistical comparisons of global severity of cognitive impairment subgroups using one-way ANOVAs, with due caution in interpretation due to potentially small group sizes and limited power.

## RESULTS

The 105 patients in OX-CHRONIC had a mean age of 68.9 years (SD = 13.0) and 40.9% were female, with a mean of 13.9 years of education (SD = 3.7), and a median acute NIHSS score of 7 (IQR =5). Further demographics and acute clinical characteristics of the cohort are outlined in Table 1. Attrition from acute to long-term follow-up was approximately 50% between each recruitment stage, primarily due to participants declining further follow-up or death (Figure 1). Full descriptive statistics of acute demographic, clinical, and cognitive characteristics across global cognitive trajectory subgroups are presented in Supplementary Table S1.

**Table 1.**
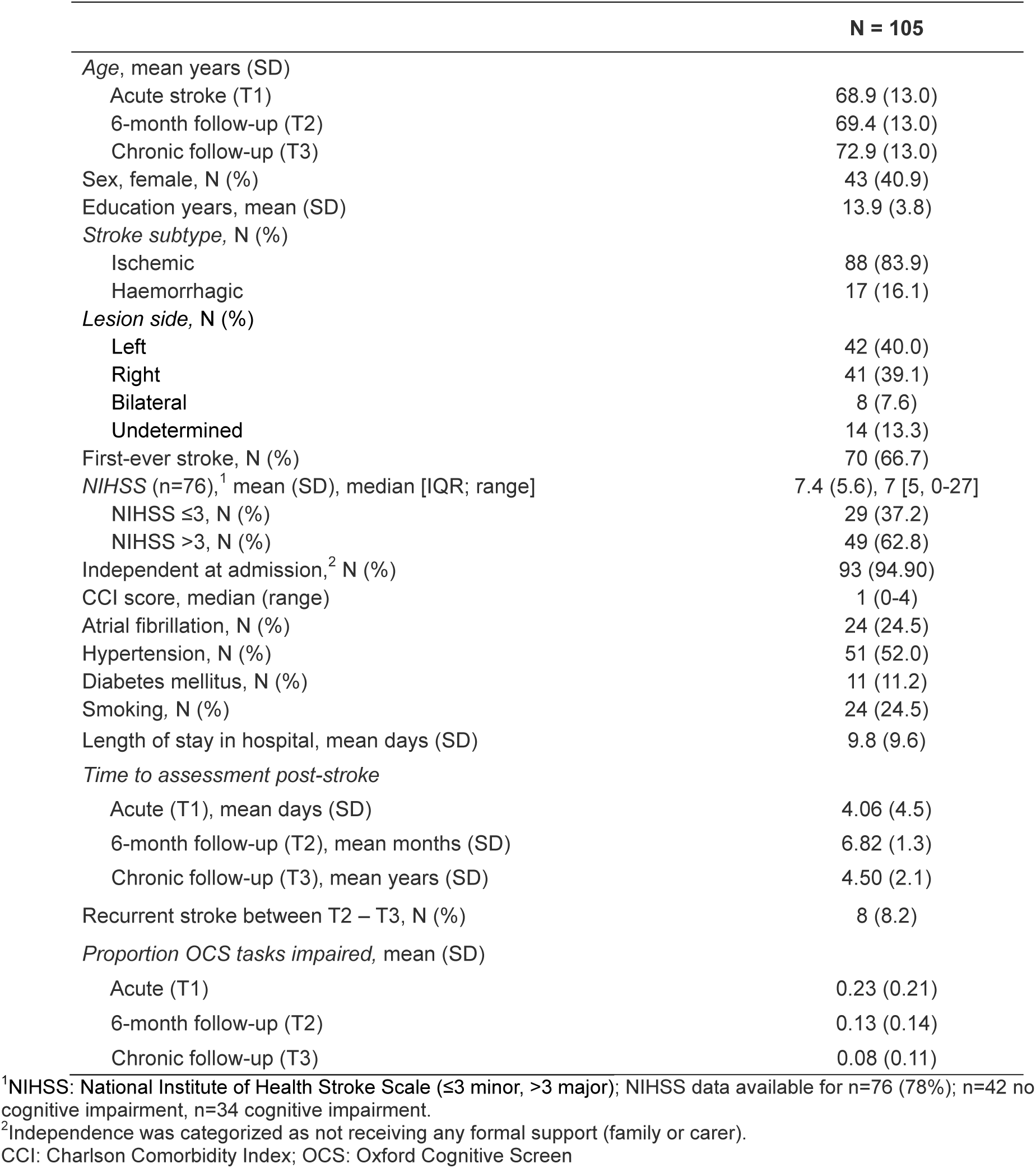
Chronic stroke cohort baseline demographics and clinical characteristics.

OCS cognitive assessments were completed at long-term follow-up (T3) in 98 patients at a mean of 4.5 years (SD = 2.1) after stroke, range 2.0–9.4 years. Details regarding prevalence of domain-specific impairments at long-term follow-up is outlined in Supplementary Table S2.

### Predicting overall severity of cognitive impairment over time

Mixed-effects models examining overall cognitive function, indexed by the proportion of OCS subtasks impaired, showed significant recovery over time (Model 1: β = −0.11 at 6 months, −0.15 at ≥2 years; *p* <0.001). In real terms, the sample on average went from having 23% of tasks impaired acutely (approximately 3 of 12 OCS tasks impaired) to on average having 12% of tasks impaired at 6-months (1-2 OCS tasks impaired), and 8% at chronic follow up (1 task impaired).

The inclusion of acute impairment status (Model 2) explained over half of the variance (*R²*=0.52), with higher acute burden predicting greater persistent deficits (*β* = 0.50, *p* <0.001). A fully adjusted model (Model 3) confirmed the predictive role of acute impairment but found no added explanatory value from demographic or clinical covariates. Random-effect variance dropped to zero once acute status was included, indicating minimal residual between-subject heterogeneity. Full model results are presented in Table 2.

**Table 2.** Predicting changes in global cognitive impairment over time (acute, 6 months, chronic follow-up).

| Global Cognitive Impairment<br>(Proportion of OCS Tasks Impaired) | *b* | SE | 95% CI | df | *t*-<br>value | Pr(>\|*t*\|) |
| --- | --- | --- | --- | --- | --- | --- |
| **Model 1** |  |  |  |  |  |  |
| (Intercept) | 0.232 | 0.021 | 0.192, 0.273 | 201 | 11.20 | <0.001*** |
| Time 2 (6 Months) | -0.107 | 0.019 | -0.143, -0.070 | 201 | -5.70 | <0.001*** |
| Time 3 (Chronic) | -0.150 | 0.020 | -0.189, -0.111 | 201 | -7.60 | <0.001*** |
| **Model 2** |  |  |  |  |  |  |
| (Intercept) | 0.115 | 0.014 | 0.088, 0.143 | 201 | 8.28 | <0.001*** |
| Time 2 (6 Months) | -0.107 | 0.017 | -0.139, -0.074 | 201 | -6.43 | <0.001*** |
| Time 3 (Chronic) | -0.155 | 0.017 | -0.188, -0.122 | 201 | -9.18 | <0.001*** |
| Acute OCS tasks impaired | 0.504 | 0.032 | 0.441, -0.568 | 103 | 15.59 | <0.001*** |
| **Model 3** |  |  |  |  |  |  |
| (Intercept) | 0.042 | 0.062 | -0.080, 0.163 | 154 | 0.67 | 0.502 |
| Time 2 (6 Months) | -0.113 | 0.019 | -0.150, -0.075 | 154 | -5.86 | <0.001*** |
| Time 3 (Chronic) | -0.181 | 0.038 | -0.255, -0.107 | 154 | -4.80 | <0.001*** |
| Acute OCS tasks impaired | 0.479 | 0.041 | 0.399, 0.560 | 69 | 11.64 | <0.001*** |
| Age | 0.0011 | 0.0007 | -0.0005, 0.0027 | 154 | 1.61 | 0.109 |
| Sex | 0.0075 | 0.016 | -0.024, 0.039 | 69 | 0.47 | 0.642 |
| Years of education | -0.0008 | 0.0023 | -0.005, 0.004 | 69 | -0.34 | 0.732 |
| Acute NIHSS | -0.0005 | 0.0014 | -0.0031, 0.0022 | 69 | -0.33 | 0.741 |
| Atrial fibrillation | -0.0039 | 0.019 | -0.042, 0.035 | 69 | -0.20 | 0.843 |
| Hypertension | -0.0126 | 0.017 | -0.045, 0.020 | 69 | -0.76 | 0.453 |
| Diabetes | 0.0046 | 0.026 | -0.047, 0.056 | 69 | 0.18 | 0.861 |
| Smoking | 0.0233 | 0.019 | -0.014, 0.061 | 69 | 1.22 | 0.228 |
| First or recurrent stroke | 0.0319 | 0.017 | -0.0019, 0.066 | 69 | 1.85 | 0.068 |
| Days post-stroke | 2.2×10<sup>-5</sup> | 2.3×10<sup>-5</sup> | -2.3×10<sup>-5</sup>, 6.6×10<sup>-5</sup> | 154 | 0.96 | 0.341 |

Three mixed-effects models were used to examine changes in global cognitive impairment over time, defined as the proportion of OCS subtasks impaired at each timepoint (acute, 6 months, and chronic follow-up). **Model 1** includes time only; **Model 2** adjusts for the severity of acute cognitive impairment (baseline OCS impairment); **Model 3** additionally includes demographic and vascular risk factors as covariates. Negative coefficients for Time 2 and Time 3 reflect a reduction in the proportion of impaired tasks compared to the acute baseline (Intercept). For dichotomous variables (atrial fibrillation, hypertension, diabetes, smoking, and recurrent stroke), values of 1 indicated presence of these variables, while 0 indicated no presence (i.e., higher beta values for first vs recurrent stroke indicate a positive association between having a recurrent stroke and global cognitive impairment).
Significance: \*\*\**p* <0.001, \*\**p* <0.01, \**p* <0.05.

### Predicting domain-specific cognitive impairment over time

Mixed-effects logistic regression models (Table 3) showed significant reductions in impairment across all domains. Memory impairment showed the greatest improvement over time, with a high odds of having improved at both post-acute follow up sessions (OR=3.65 at 6 months, 16.40 at ≥2 years). However, having an acute impairment substantially increased long-term risk (OR=55.2, 21.5–141.0; mR²=0.60).

**Table 3.** Predicting changes in domain-specific cognitive impairments over time (acute, 6 months, chronic/long-term follow-up).

| *Domain-specific impairment* | Log odds | OR | SE | 95% CI | z-value | Pr(>\|z\|) |
| --- | --- | --- | --- | --- | --- | --- |
| **Language** |  |  |  |  |  |  |
| (Intercept) | -1.314 | 0.27 | 0.355 | [0.13, 0.54] | -3.701 | <0.001 *** |
| Time 2 (6 Months) | 1.245 | 3.47 | 0.430 | [1.50, 8.07] | 2.896 | 0.004 ** |
| Time 3 (Chronic) | 2.101 | 8.17 | 0.483 | [3.17, 21.1] | 4.346 | <0.001 *** |
| Acute Language Impaired | 3.399 | 29.9 | 0.413 | [13.3, 67.2] | -8.237 | <0.001 *** |
| *Language: mR<sup>2</sup> = 0.506, ICC<sub>adj</sub> = 0.00* |  |  |  |  |  |  |
| **Memory** |  |  |  |  |  |  |
| (Intercept) | -1.635 | 0.20 | 0.400 | [0.09, 0.43] | -4.088 | <0.001 *** |
| Time 2 (6 Months) | 1.271 | 3.56 | 0.472 | [1.41, 8.99] | 2.693 | 0.007 ** |
| Time 3 (Chronic) | 2.797 | 16.40 | 0.555 | [5.52, 48.7] | 5.038 | <0.001 *** |
| Acute Memory Impaired | 4.010 | 55.20 | 0.480 | [21.5, 141.0] | 8.359 | <0.001 *** |
| *Memory: mR<sup>2</sup> = 0.601, ICC<sub>adj</sub> = 0.00* |  |  |  |  |  |  |
| **Attention** |  |  |  |  |  |  |
| (Intercept) | -1.336 | 0.26 | 0.299 | [0.15, 0.47] | -4.465 | <0.001 *** |
| Time 2 (6 Months) | 1.295 | 3.65 | 0.359 | [1.81, 7.38] | 3.607 | <0.001 *** |
| Time 3 (Chronic) | 1.688 | 5.41 | 0.381 | [2.57, 11.4] | 4.434 | <0.001 *** |
| Acute Attention Impaired | 2.409 | 11.10 | 0.363 | [5.46, 22.6] | 6.644 | <0.001 *** |
| *Attention: mR<sup>2</sup> = 0.369, ICC<sub>adj</sub> = 0.02* |  |  |  |  |  |  |
| **Executive function** |  |  |  |  |  |  |
| (Intercept) | -0.783 | 0.46 | 0.355 | [0.23, 0.92] | -2.205 | 0.027 * |
| Time 2 (6 Months) | 1.051 | 2.86 | 0.439 | [1.21, 6.77] | 2.389 | 0.017 * |
| Time 3 (Chronic) | 1.421 | 4.14 | 0.462 | [1.68, 10.2] | 3.078 | 0.002 ** |
| Acute Executive Impaired | 2.886 | 17.90 | 0.389 | [8.36, 38.40] | 7.420 | <0.001 *** |
| *Executive function: mR<sup>2</sup> = 0.377, ICC<sub>adj</sub> = 0* |  |  |  |  |  |  |
| **Number processing** |  |  |  |  |  |  |
| (Intercept) | -1.404 | 0.25 | 0.360 | [0.12, 0.51] | -3.736 | <0.001 *** |
| Time 2 (6 Months) | 2.457 | 11.7 | 0.504 | [4.35, 31.4] | 4.876 | <0.001 *** |
| Time 3 (Chronic) | 2.012 | 7.48 | 0.482 | [2.91, 19.4] | 4.176 | <0.001 *** |
| Acute Number Impaired | 3.523 | 33.99 | 0.467 | [13.6, 84.6] | 7.548 | <0.001 *** |
| *Number processing: mR<sup>2</sup> = 0.550, ICC<sub>adj</sub> = 0* |  |  |  |  |  |  |

Mixed-effects logistic regression models for each cognitive domain. The table shows log odds, odds ratios (OR), standard errors (SE), 95% confidence intervals (CI), z-values, and p-values for the fixed effects. Model performance metrics include marginal R<sup>2</sup> (mR<sup>2</sup>) and adjusted intraclass correlation coefficient (ICC<sub>adj</sub>). In models where singular fits were detected (i.e., random effect variance ≈ 0), ICC<sub>adj</sub> is not reported. Significance levels: \*\**p* < 0.001, \**p* < 0.01, *p* < 0.05.

Similarly, language abilities also improved at 6 months (OR=3.47, 95% CI 1.50–8.07; *p*=0.004) and further at ≥2 years (OR=8.17, 3.17–21.1; *p*<0.001), with acute impairment strongly associated with persistent deficits at follow-up (OR=29.9, 13.3–67.2; mR²=0.51). Attention impairment improved over time (OR=3.65 at 6 months, 5.41 at ≥2 years), with acute deficits remaining a strong predictor of persistence (OR=11.1, 5.46–22.6; mR²=0.37). Executive-function impairment also improved (OR=2.86 at 6 months, 4.14 at ≥2 years), with acute impairment linked to a 17-fold increase in long-term risk (OR=17.9, 8.36–38.40; mR²=0.37). Number-processing impairment showed the steepest early improvement (OR=11.70 at 6 months), which was sustained at follow-up (OR=7.48, 2.91–19.4); similarly, acute impairment increased odds of chronic impairment (OR=34.99, 13.6–84.6; mR²=0.55). Across models, random-effect variance was negligible (ICCadj ≤0.02 or singular), indicating minimal residual between-subject variability after accounting for time and baseline status.

### Trajectories of global cognitive impairment

In the global cognitive impairment model, the four-class model demonstrated the best balance between fit, parsimony, and interpretability (AIC = −371.73, BIC = −329.27, SABIC = −379.82, H = 0.80; Table 4). Although the two-class solution had better entropy (i.e., class separation), it was judged to oversimplify the heterogeneity observed in post-stroke cognitive recovery profiles. The final model identified four distinct trajectories (Figure 2):

- Class 1: No or mild acute impairment with stable performance over time (47.62%);
- Class 2: Moderate initial impairment with improvement over time (32.38%);
- Class 3: Severe impairment with large improvement over time (15.24%);
- Class 4: Moderate acute impairment with decline (4.76%).

**Figure 2.**
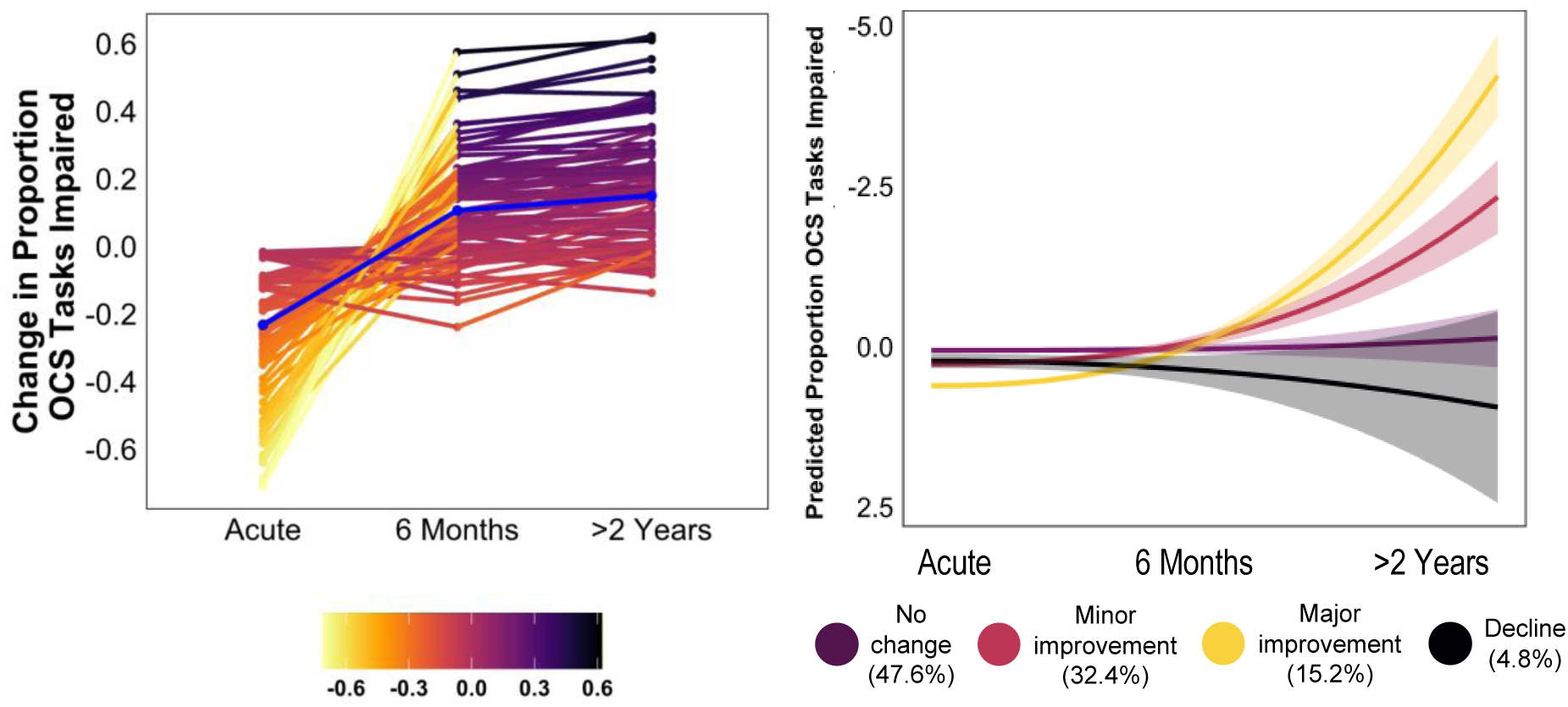
Trajectories of global cognitive impairment following stroke. Distinct trajectories of post-stroke global cognitive impairment over 3 timepoints (acute, 6 months, chronic follow-up) for each of the four classes identified by Latent Class Growth Modeling. Global cognitive impairment was defined by the proportion of OCS tasks impaired. Classes are labeled by colour and percentage of participants assigned to each latent class is displayed. Left-side: individual change in the raw variable (proportion OCS tasks impaired) over time, with positive change indicating improvement (in darker colours); Right-side: shows predicted proportion of OCS tasks impaired over time per identified latent class identified.

**Table 4.** Model fit statistics and class membership for Latent Class Growth Analysis of Global Cognitive Impairment.

| Number of classes | Free parameters | Log Likelihood | AIC | BIC | SABIC | H | Class Membership (%) |  |  |  |  |
| --- | --- | --- | --- | --- | --- | --- | --- | --- | --- | --- | --- |
|  |  |  |  |  |  |  | 1 | 2 | 3 | 4 | 5 |
| 1-class | 4 | 127.38 | -246.76 | -236.15 | -248.78 | 1.00 | 100.00 |  |  |  |  |
| 2-class | 8 | 187.88 | -359.76 | -338.53 | -363.80 | 0.94 | 80.95 | 19.05 |  |  |  |
| 3-class | 12 | 192.51 | -361.01 | -329.17 | -367.08 | 0.70 | 17.14 | 31.43 | 51.43 |  |  |
| **4-class** | **16** | **201.87** | **-371.73** | **-329.27** | **-379.82** | **0.80** | **47.62** | **32.38** | **15.24** | **4.76** |  |
| 5-class | 20 | 201.87 | -363.73 | -310.65 | -373.84 | 0.63 | 15.24 | 0.00 | 40.00 | 4.76 | 40.00 |

AIC: The Akaike Information Criterion, a measure of model fit that penalizes for the number of parameters; BIC: The Bayesian Information Criterion, another measure of model fit that penalizes more heavily for the number of parameters; SABIC: Sample-Adjusted BIC; H: Entropy; Measure of class separation; Profile Membership (%): The percentage of participants assigned to each latent class.

Full model fit statistics and class membership distributions are shown in Supplementary Table S3. As expected, classes differed on baseline cognitive severity (greater proportion of OCS tasks impaired) and total number of domains impaired. Chronic MoCA scores also differed across classes (F=12.67, *p* <0.001), with lower scores observed in the declining (M = 19.3 [SD=5.8]) and persistently impaired (M = 19.1 [SD=3.5]) groups compared to those with moderate acute impairment (M = 23.3 [SD=3.1]) and either stable or mild impairment (M = 24.8 [SD=3.5]; Supplementary Table S1).

### Acute Profiles of Global Cognitive Impairment Subgroups

Full descriptive statistics of acute demographic, clinical, and cognitive characteristics across global cognitive trajectory subgroups are presented in Supplementary Table S1. Participants with stable mild or no impairments post-stroke had more years of education, smaller lesion volumes, and were less likely to show multi-domain impairment in the acute phase. The declining subgroup showed lowest education levels and, along with those with severe acute impairments, showed the lowest MoCA scores at long-term follow-up (M = 19.3 [SD=5.8]).

### Trajectories of domain-specific cognitive impairments

Latent class trajectories for each cognitive domain are visualised in Figure 3, with model fit statistics reported in Supplementary Tables S3–7. A 3-class solution best fit the data for language (AIC = −273.83, BIC = −241.99, SABIC = −279.89, H = 0.99), memory (AIC = −257.75, BIC = −225.90, SABIC = −263.81, H = 0.97), number processing (AIC = −110.98, BIC = −79.13, SABIC = −117.04, H = 0.98), and executive function (AIC = 1525.28, BIC = 1557.13, SABIC = 1519.22, H = 0.90). For attention, a 4-class model provided the best fit (AIC = −29.56, BIC = 12.91, SABIC = −37.64, H = 0.92). Across domains, some classes showed similar patterns (e.g., late improvements from 6-months to long-term follow-up), while executive function uniquely included a subgroup with fluctuating performance over time. Individual-level raw trajectories for each domain are presented in Supplementary Figure S1 and illustrate the within-class variability underlying these modelled patterns.

**Figure 3.**
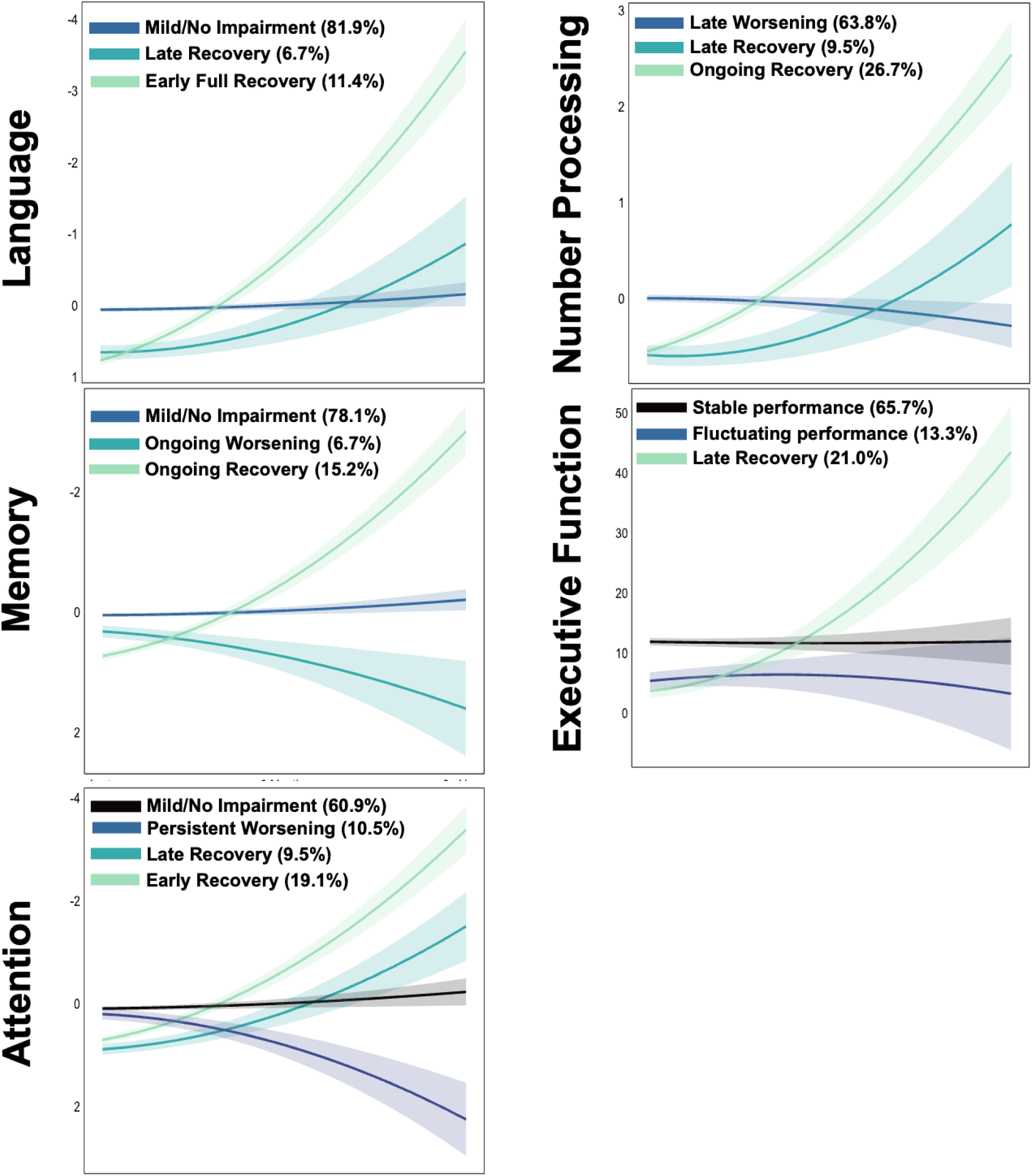
Trajectories of domain-specific cognitive impairments following stroke. Distinct trajectories of domain-specific cognitive impairment across three timepoints (acute, 6 months, and chronic follow-up), identified using Latent Class Growth Modeling. For the language, memory, attention, and number processing domains, impairment was defined by the proportion of domain-specific subtasks impaired. For executive function, raw scores from the mixed trails subtask were used. Each plot displays predicted class-level trajectories, with class labels and the percentage of participants in each class shown. The y-axis represents the predicted level of impairment over time.

## DISCUSSION

This study characterized global and domain-specific cognitive trajectories up to 9 years post-stroke (mean follow-up 4.5 years) using repeated, domain-sensitive assessments. In the majority of individuals, both global and domain-specific impairments declined over time, with the greatest recovery occurring within the first six months and continued improvement observed in some domains, particularly memory and language. Acute cognitive impairment was the strongest and most consistent predictor of long-term outcomes, while demographic and vascular factors explained minimal additional variance. Importantly, latent class analyses revealed substantial heterogeneity in recovery patterns, including subgroups with delayed improvement or late decline across all domains, highlighting that one-off assessments may fail to capture critical change. These findings underscore the need for individualized monitoring of long-term cognitive outcomes, information provision on possible post-stroke trajectories, and customising cognitive support and rehabilitation based on individual trajectories observed. Future research efforts should aim to replicate trajectories found here and better characterise baseline features using a range of methods so as to better predict recovery trajectories. By identifying replicable markers of trajectories of improvement, persistent impairment and decline, rehabilitation efforts can be targeted more appropriately.

### Global cognitive impairment

A significant reduction in global cognitive impairment over time was observed, with the steepest improvements in function occurring within the first six months post-stroke and further, though attenuated, gains evident at long-term follow-up (mean 4.5 years, range 2–9 years). Mixed-effects modelling showed a consistent reduction in the proportion of OCS tasks impaired, going from on average 3 OCS subtasks impaired at baseline to 1-2 OCS subtasks impaired by the 6-month and chronic phase These findings align with prior studies demonstrating early subacute cognitive recovery (25) but extend them by showing sustained improvement well beyond the assumed one-year plateau (26), and support data from longitudinal cohorts showing recovery up to two (27) and even ten years post-stroke (8, 13). This long-term improvement may reflect a combination of rehabilitation efforts, spontaneous restitution, behavioural compensation, and persistent neuroplasticity (28, 29). Of note, this may only be true of those assessed here, given those with worsening impairments withdrew from our sample or passed away.

The strongest and most consistent predictor of global cognitive outcome was the severity of acute cognitive impairment, which accounted for over half the variance in performance across timepoints (R²=0.52). Once acute cognitive burden was included, additional demographic, vascular, and clinical variables contributed negligible explanatory power, mirroring meta-analytic findings (17), and domain-level studies showing acute deficits to be the strongest predictors of longer-term impairment (2, 9, 30). This suggests that early multi-domain cognitive screening may serve as a proxy for global cerebral vulnerability at the time of stroke, incorporating both acute lesion burden and pre-existing pathology (9, 31). These findings strongly support the clinical utility of acute cognitive profiling and its inclusion in early stroke care to inform prognostication, discharge planning, and resource allocation, an approach consistent with current guideline recommendations (7, 32).

However, our findings contrast to recent large-scale pooled data from Lo et al. (12), who reported an acute drop in global cognition followed by an accelerated decline in over 20,000 community-dwelling older adults, with no evidence of post-stroke recovery. Several methodological differences may explain this divergence. While Lo et al. (12) harmonized disparate screening tools across cohorts, we used a validated, domain-specific measure (OCS) with high sensitivity to stroke-related cognitive deficits and the ability to distinguish them from sensorimotor symptoms. In addition, our hospital-verified cohort underwent lesion and cognitive phenotyping, and our analytic approach explicitly modelled heterogeneity using latent class growth analysis, rather than assuming a single mean trajectory. Finally, our cohort predominantly included moderate strokes (mean NIHSS=7), with a majority showing focal cognitive impairments, whereas the majority of studies on post-stroke cognition are biased towards milder stroke samples with lesser opportunity for recovery.

Latent class analysis identified four distinct global cognitive recovery patterns. Nearly half the cohort (47.6%) exhibited stable mild or no impairment throughout follow-up. A further 32.4% presented with moderate impairment acutely but improved substantially, and 15.2% showed severe acute deficits with notable long-term gains. In contrast, 4.8% experienced delayed cognitive decline following an initially moderate presentation. This latter class resembles the “plateau-then-decline” trajectory observed by Delgado et al. (33) and may reflect the impact of progressive small vessel disease (SVD), covert recurrent infarction, or emerging neurodegeneration (31, 34).

The clinical implications are considerable. First, these findings support early cognitive screening as a tool for risk stratification and long-term care planning. Second, the substantial recovery seen in individuals with moderate or even severe acute deficits reinforces the potential for ongoing cognitive rehabilitation to maximise improvements made over time. This research also challenges deterministic models of post-stroke decline. Finally, the identification of a small but meaningful subgroup with delayed deterioration highlights the need for routine, long-term cognitive monitoring, ideally embedded within a stroke care pathway, to detect emerging decline and intervene appropriately.

### Domain-specific cognitive impairment

Unlike global scores, which may obscure important heterogeneity, domain-level modelling captured differential trajectories that reflect the neuroanatomical and functional specificity of stroke-related damage. Mixed-effects models showed significant reductions in impairment over time across all domains assessed, including language, memory, attention, executive function, and number processing, with the most pronounced recovery typically observed within the first six months. However, the degree and persistence of recovery differed by domain.

### Language

Language function showed the most favourable recovery profile across domains. Latent class modelling identified three distinct trajectories: the majority (81.9%) exhibited no or only mild impairment throughout follow-up; 11.4% showed early full recovery; while 6.7% showed late improvements. These patterns are consistent with prior studies showing rapid recovery from aphasia within the first three months post-stroke, often attributed to spontaneous neurobiological restitution (35) However, evidence from longitudinal cohorts suggests that meaningful gains can continue well into the chronic phase, likely supported by adaptive functional reorganization and therapy-driven gains (36).

### Memory

Memory recovery showed a broadly favourable profile, though less consistent than language, with a greater proportion of individuals showing persistent impairment or delayed improvement. Latent class modelling identified three trajectories: no or mild acute impairment that remained stable (78.1%), ongoing recovery (15.2%) and ongoing worsening (6.7%). These findings align with previous studies showing early memory gains post-stroke, alongside considerable long-term variability (37, 38). Acute memory impairment remained a strong predictor of long-term outcome, consistent with studies linking early deficits to persistent dysfunction (39), though we note that memory was also the most well-recovered domain, potentially linked to language recovery and therefore improved verbal encoding. The observed heterogeneity in recovery may reflect the distributed neuroanatomical basis of memory, encompassing medial temporal, prefrontal, and subcortical structures variably affected by stroke and SVD (37, 40).

### Attention

Attentional recovery was more variable than language and memory, with a notable minority of patients exhibiting persistent or worsening deficits. Four trajectory patterns emerged: no or mild impairment across follow-up (60.9%), early recovery (19.1%), delayed recovery (9.5%), and progressive decline (10.5%).

These findings confirm substantial heterogeneity often masked by global cognitive scores and align with prior studies showing persistent attention deficits in many stroke survivors, even years after the index event (41, 42). Persistent impairments in attention are particularly concerning given their association with reduced independence, increased safety risks, and poorer performance across other cognitive domains, including executive function and memory encoding (2, 16).

### Executive function

Executive function showed the least favourable recovery profile. Three trajectories were identified: stable performance (65.7), fluctuating performance (13.3%), and late recovery (21.0%). Mixed-effects modelling confirmed only a modest reduction in impairment over time (OR=0.53, i.e. a 47% reduction in the odds of full recovery), and acute impairment remained the strongest predictor of long-term outcome. Therefore, a sizeable subgroup experiences executive dysfunction that persists or worsens, with limited spontaneous recovery. This pattern mirrors prior work in both mixed-age and young-stroke cohorts(43, 44), where only 20% showed meaningful gains at one year, while the majority remained stable or declined (10). A systematic review further highlights the variability in executive recovery, with many studies failing to demonstrate improvements beyond measurement noise (6).

Persistent executive dysfunction may reflect the vulnerability of this domain to both focal and diffuse pathology. Executive control relies on distributed frontostriatal and frontoparietal networks, which are particularly sensitive to the effects of white matter disconnection and SVD (45). Moreover, persistent executive impairment has been identified as a key predictor of post-stroke dementia, particularly in the presence of mixed vascular and neurodegenerative pathology (16, 33).

### Number processing

Number processing recovery showed considerable heterogeneity. The majority (57.1%) showed stable mild/no impairment, 23.8% ongoing improvement, 9.5% late improvement, and 9.5% experienced delayed decline. The variability in recovery patterns likely reflects the complex neurocognitive demands of numerical cognition, which rely on distributed frontoparietal and subcortical networks susceptible to both focal and diffuse brain injury (46, 47).

### Strengths and limitations

This study has several strengths. A key advantage is the long-term, domain-specific cognitive follow-up extending up to 9.3 years post-stroke, offering insights into the chronic course of cognitive trajectories. The inclusion of moderate-to-severe stroke survivors and individuals with aphasia, groups often excluded from research (48), enhances the generalizability of our findings. Use of the stroke-specific OCS enabled sensitive detection of both overall cognitive impairment severity and domain-level impairments, allowing us to characterise distinct recovery trajectories across cognitive domains. However, various limitations should be acknowledged. Despite best efforts, attrition over time, largely due to death or severe illness, was considerable, as is common in long-term stroke studies, especially in older cohorts. This may have led to underestimation of cognitive impairment and decline, as individuals lost to follow-up likely had more severe deterioration (49) and cognitive trajectories in more impaired patients therefore remains uncertain. A further limitation is the absence of pre-stroke cognitive data, due to recruitment during acute hospitalization. This restricts our ability to disentangle pre-existing cognitive decline from stroke effects.

Prior studies suggest that individuals who go on to experience stroke may already exhibit accelerated cognitive decline up to a decade before the event (50). Further, though exploratory analyses between overall cognitive severity subgroups is valuable to guide future prediction of those at risk of decline, some subgroups constituted relatively small numbers of participants, affecting power. Additionally, due to the remote nature of follow-up, the praxis domain could not be assessed beyond 6 months, limiting insights into long-term praxis recovery. Finally, while our modelling accounted for a range of clinical and demographic factors, remaining unexplained variance suggests unmeasured factors, such as rehabilitation intensity, social engagement, or genetic vulnerability, may have contributed to changes in long-term cognition.

Future work should aim to replicate our findings in larger and more diverse samples, and where feasible, incorporate pre-stroke cognitive measures (e.g., through proxy measures of cognition such as the IQ-CODE), though these remain challenging to obtain. Longitudinal studies that combine early cognitive phenotyping with detailed neuroimaging may further clarify the mechanisms driving divergent recovery trajectories. In addition, further work is required to enable individualised prediction of specific cognitive trajectories to enable targeted follow-up in those at greatest risk of long-term cognitive decline and to provide reassurance regarding the high likelihood of spontaneous improvement. Finally, examining whether personalised, trajectory-informed interventions improve outcomes relative to standard care represents an important next step for clinical application.

## Conclusion

This study provides rare longitudinal insight into long-term post-stroke cognitive recovery, revealing that global and domain-specific functions often improve well beyond the subacute phase, particularly in language, memory, and number processing. Acute cognitive impairment emerged as the strongest and most consistent predictor of long-term outcomes, while demographic and vascular factors added limited explanatory value. Distinct trajectories across all domains, ranging from early and delayed recovery to persistent deficits and late decline, underscore the need to move beyond static models of cognitive outcome. In particular, executive dysfunction and attention deficits were less amenable to spontaneous recovery. These findings highlight the importance of early, domain-sensitive screening to inform prognosis and rehabilitation, while supporting a shift toward dynamic, individualized cognitive surveillance.

Incorporating trajectory-informed monitoring into stroke care may improve the identification of individuals at risk for decline and guide resource allocation, a strategy that merits prospective evaluation in future trials to determine whether trajectory-based stratification improves functional and cognitive outcomes.

## Data Availability

The Data has been made freely accessible through Dementias Platform UK and can be accessed at www.dementiasplatform.uk

## ACKNOWLEDGMENTS

We thank all participants and members of the Oxford Translational Neuropsychology Group for contributions to recruitment/testing.

